# Evaluation of the Clinical Performance of Two New Onchocerciasis Rapid Diagnostic Tests

**DOI:** 10.1101/2025.09.24.25336608

**Authors:** Dziedzom K. de Souza, Yvonne Ashong, Selassie Afatodzie, Mawunyo Dogbe, Abena Akyeamaa Nyarko, Bernard Logonia, Kwadwo Frempong, Carson Moore, Lee Hundley, Philip Kutjok, Jessica Prince-Guerra, Scott E. Elder, Kimberly Y. Won, Christophe Vedrine, Philippe Leissner, Annelyse Duvoix, Emily Adams, Marco Biamonte, Daniel A. Boakye

## Abstract

Current diagnostic methods for onchocerciasis lack sufficient utility, sensitivity or specificity. Two new lateral flow assays developed by Bioaster and Global Access Diagnostics (BA/GADx) and Drugs and Diagnostics for Tropical Diseases (DDTD) multi-antigen test Biplex Version C (DDTDc) were evaluated in Ghanaian communities to assess their potential for point-of-care use. The study enrolled 1,700 participants (≥5 years) across seven communities. The tests were evaluated on whole blood in the field and dried blood spots (DBS) in the laboratory. The results were compared with the existing SD Ov16 RDT test of DBS and O-150 qPCR on skin snips. Cross-reactivity was assessed via *Mansonella perstans* qPCR. The sensitivities of the DDTDc compared to the SD Ov16 ranged from 75.52% - 85.57%, while that of the BA/GADx ranged from 83.33% - 88.74%. The confidence intervals for the sensitivity of these two tests overlap. Tests on the DBS yielded the highest sensitivity for both tests. Skin snip qPCR identified only 26 positives, limiting specificity assessments. Both tests demonstrated field usability and adequate sensitivity compared to the SD Ov16 RDT. These advancements represent progress toward diagnostic tools available for elimination programs but underscore the need for continued diagnostic innovation to address the lingering challenges.

## Introduction/Rationale

Human onchocerciasis, commonly known as “river blindness”, is a neglected tropical disease caused by the filarial nematode *Onchocerca volvulus*. It is the world’s second leading cause of infectious blindness after trachoma [1]. The parasite is transmitted by blackflies of the genus *Simulium* through repeated bites where the infective larvae (L3) are inoculated into the human host. The presence and the death of microfilariae (mf) in the skin and eyes are associated with severe skin itching, epilepsy, intellectual disabilities, visual impairment and permanent blindness [1, 2]. Onchocerciasis affects about 20.9 million people worldwide, with 99% of cases found in 31 countries in sub-Saharan Africa. It is primarily combatted through mass drug administration (MDA) of ivermectin, a deworming drug. However, limitations in diagnostic tools capable of detecting infections [3, 4] and confirming disease presence/absence within endemic regions has hampered the ability to confirm elimination of the disease.

The reference method for diagnosis of onchocerciasis is identification of microfilariae (mf) by microscopic examination of skin snips from the iliac crest or by molecular detection methods [5]. While these techniques have good specificity, they have low sensitivity for early-stage infections and in areas with low infection levels. The tests most widely used by programs detect antibodies to the recombinant antigen Ov16 and are available in multiple formats, including ELISA and lateral flow assays, often referred to as rapid diagnostic tests (RDT). The commonly used Ov16 ELISAs have reported sensitivities of 80% - 88.2% and specificities of 97% - 99.98% [6–8]. Another antibody test, the SD Bioline Ov16 RDT, is currently used for monitoring and surveillance in onchocerciasis programmes [6]. However, its current usage is recommended on eluted dried bloodspots (DBS) in the laboratory and not on whole blood in the field [9]. A drawback to antibody tests is the inability to distinguish current infection from past exposure and infection. In addition, given the performance of existing tests, there are limitations in measuring current programmatic serological thresholds of 0.1% for stopping MDA [10] and the 2% anti-Ov16 prevalence in adults for starting MDA [11]. Despite these limitations, antibody tests have been used in mapping hypo-endemic areas and detecting recent transmission in children [12].

The WHO Onchocerciasis Technical Subgroup (OTS) has recognized the need for improved diagnostic tools to use in low-prevalence settings. To address this need, the WHO Diagnostic Technical Advisory Group (DTAG) onchocerciasis sub-group identified onchocerciasis elimination mapping (OEM) and MDA stopping as priority use cases for diagnostics development. Thus, the DTAG developed target product profiles (TPPs) for diagnostics to use in these program stages, each with distinct performance characteristic requirements [13]. The TPP for OEM recommends a sensitivity of ≥60%, while the TPP for MDA stopping decisions suggests a sensitivity of ≥89%. Both TPPs set an ambitious goal of ≥99.8% specificity. New point of care (POC) diagnostics aimed at meeting these programmatic needs have been developed recently and have undergone laboratory evaluations.

These new tests include the Bioaster and Global Access to Diagnostics (BA/GADx) test and Drugs and Diagnostics for Tropical Diseaseas (DDTD) test. The DDTD test is a biplex RDT with two test lines, configured to detect IgG4 antibodies to Ov16 at test line 1 (T1) and to OvOC3261 at test line 2 (T2). The Biplex C (referred in this paper as DDTDc) is a simplification and an improvement over a previous version (Biplex A) which contained these and two other antigens and was previously evaluated in South Sudan [14]. The BA/GADx detects IgG4 antibodies against the *O. volvulus*-specific antigen, Ov16 and is similar to the SD Bioline Ov16 RDT [15]. Both tests are designed to provide rapid results directly at POC. This study aimed to generate data on a head-to-head clinical performance of these new diagnostics when used at POC in both field and laboratory settings. The study also assessed the ease-of-use of the tests.

## Methods

### Study sites

This study was conducted in three districts in Ghana, where onchocerciasis control activities have been ongoing since the 1970s [16]. The field evaluation of the new tests took place in seven rural communities with varying levels of onchocerciasis endemicity. The sites were selected to represent areas of moderate to high prevalence, low prevalence, and non-endemic areas. Thus, we identified sites where field data (unpublished) existed to indicate the prevalence of onchocerciasis (**Table 1**). For assessments of specificity, sites were also selected based on a probability of identifying co-endemic infections (lymphatic filariasis (LF), mansonellosis) and, ideally, a low prevalence of onchocerciasis.

### Target population and sample size estimation

The target population for this study was individuals ≥5 years, with efforts to enroll children and adults with approximately equal numbers of males and females. The sample size was calculated to assess non-inferiority of sensitivity of the DDTDc and BA/GADx RDTs compared to the SD Ov16 DBS on RDT. The primary outcome was the percentage of participants testing positive by each RDT, and these percentages were compared across the different test types.

The null hypothesis (H_0_) was that the sensitivity of the SD Ov16 RDT (π□) exceeded that of the DDTDc or BA/GADx tests (π_e_) by a clinically relevant margin (d): **H**_**0**_**: π** □ **≥ π**_**e**_ **+ d**. The alternative hypothesis (H_1_) was that the sensitivity of the DDTDc or BA/GADx RDTs was not inferior by more than this margin: **H**_**1**_**: π**□ **− d < π**_**e**_. A non-inferiority margin of 1% (d = 0.01) was chosen to reflect the smallest difference considered clinically meaningful. Assuming a true positivity rate of 1% for all test groups, a one-sided alpha of 5%, and 90% power, the required sample size was 1,697 participants. This was rounded up to 1,700 to ensure adequate power. Sample size calculations were performed using the Sealed Envelope online calculator for binary non-inferiority trials (Sealed Envelope Ltd. 2012; https://www.sealedenvelope.com/power/binary-noninferior/; accessed May 16, 2023).

### Sampling strategy

All study personnel were trained to ensure that all protocols and diagnostic test procedures were understood and followed. The training included a theoretical and field practice component. Pre-test and post-test evaluations were undertaken by the study personnel to evaluate the increase in knowledge among trainees after training.

Study Participants were recruited prospectively until the sample size was met. At least 242 participants were targeted per site, except in Gborsike and Lancha where the population size was comparatively smaller (see Table 6). In each selected site, a convenience sample of individuals ≥ 5 years old was selected for specimen collection. During the survey, the study team arrived at the pre-selected community either on the eve of the survey date or early in the morning on the day of the survey and proceeded to brief the stakeholders. In each selected site, community entry procedures were followed as appropriate to obtain permission from the chiefs and opinion leaders. Community mobilization was undertaken in each of the study communities, through durbars, and using the community radio stations. The team proceeded to a location offered by the community (health centre, schools, etc.) to carry out the survey. Participants were enrolled by convenience sampling – i.e. people were enrolled in the study as they came to the survey site. Written informed consent was collected from adult, and parental consent and child assent for children aged 12-17 years. A questionnaire was administered to participants to collect demographic information, onchocerciasis related knowledge, attitude and practices, and willingness and ability to provide the required biological samples. In each community, the survey team spent up to 5 days to reach the maximum number of people.

### Specimen collection

Blood samples were collected from community members who agreed to participate in the study. A maximum of 7.5 mL of EDTA blood was collected to test for the BA/GADx, DDTDc, SD Ov16 RDT, LF FTS (only in LF co-endemic areas), *M. perstans* qPCR, *W. bancrofti* microscopy, preparation of dried blood spots (DBS), repeat tests and storage (based on the participant’s consent) (**Table 2**). Two skin snips were collected for adult participants for skin-snip microscopy and qPCR.

### Fingerstick blood collection and RDT procedures in the field

In the communities, the field team tested participants with the DDTDc and BA/GADX RDTs. as well as the FTS for LF (Adaklu only). The field tests were conducted with blood collected directly from a finger using the collection device provided by the manufacturer and applied directly to the RDT. All tests were conducted based on manufacturer’s instructions for use. Approximately 100ul of blood was used for the three tests (see **Table 2** for the volume per test). The results for each test were read at the time indicated in the manufacturer’s guidance. Laboratory tests that were invalid or had indeterminate results were repeated once. All results, including invalid or indeterminate results, were recorded using the electronic data collection system developed for the study. The test result was also noted on the test cassette, indicating the time of reading, and a picture was taken of all tests. The pictures were linked to each participant’s results in the electronic data collection system.

### Venous blood collection for the preparation of dried blood spots (DBS), Mansonella perstans qPCR and storage

Approximately, 6.0 mL of whole blood was collected from participants into EDTA vacutainer tubes. DBS were prepared for each participant for subsequent laboratory-based testing with the BA/GADx and the DDTDc tests (based on manufacturer’s instructions for use). The SD Ov16 RDT was performed in the laboratory on DBS in line with WHO recommendations [9]. The DBS was also used to test for *M. perstans* using qPCR methodology [17]. Two TropBio filter paper disks were made per subject to provide up to twelve 10 μL blood spots. The filter disk was allowed to air dry completely before storing (-20 °C) and transporting to the laboratory. Two 2ml aliquots of blood were prepared and the remaining blood sample was centrifuged to separate plasma and red cells and stored.

### Skin snips

The skin snip biopsy was performed using a sterile sclerocorneal biopsy punch after appropriate disinfection of the skin by an experienced technician. Two snips were taken for consenting participants 18 years and above. The snips were then incubated in normal saline at room temperature for 24 hours to allow the mf to emerge. The mf were counted and the skin snips and saline solution transferred into a 2 mL Eppendorf tube for subsequent testing using the O-150 qPCR [18].

### Night blood collection and thick blood smear procedure for W. bancrofti

Due to the nocturnal periodicity of *W. bancrofti)*, any individuals positive by FTS were followed up for a second blood sample collection from 21:00 – 24:00 for the preparation of thick blood smear, and assessment of mf loads.

### Feasibility and Ease of Use Assessment

The feasibility and ease-of-use tests were assessed by administering a questionnaire (See Supplementary File 1) to four test readers, who consistently carried out the tests throughout the study.

### *Data management* and analysis

An Electronic Data Collection (EDC) form was developed for the study using REDCap. The data were collected on Samsung Galaxy Tab S2 tablets and uploaded regularly to an online database. The study personnel were trained in using these tools for data collection. QR code labels were printed for all participants and sample types collected. In addition to the use of the EDC, all test results were recorded in field or laboratory notebooks.

The Kappa coefficient was calculated comparing the percent agreement between the SD Ov16 RDT and each of the new tests, and between novel diagnostics. Sensitivity and specificity were calculated using both skin snip O-150 PCR and the SD Ov16 RDT as reference standards. The data were tabulated and analysed using MedCalc online and GraphPad Prism (10.4.2). All prevalence estimates were accompanied by the 95% confidence intervals. Comparison between age groups was done using the one-sample t-test, with significance set at *p* = 0.05.

### Ethical Considerations

This study (CPN 013/23-24) was reviewed and approved by the Institutional Review Board of the Noguchi Memorial Institute for Medical Research, University of Ghana, with Federal Wide Assurance FWA 0001824. Written informed consent was obtained from all adult participants. For children 12 years and above, written assent was obtained in addition to parental consent. For children under 12 years, only parental consent was obtained. CDC reviewed the protocol and determined CDC to be non-engaged in the conduct of this research – see the Code of Federal Regulations: 45 C.F.R. part [19]; 21 C.F.R. part 56 [20].

## Results

### Participant recruitment and demographics

From September 24, 2023, to October 20, 2023, 1913 participants were consented, out of which 1700 were finally enrolled into the study. Consenting was done in participants’ place of residence and only those who reported to the sample collection point were finally enrolled until the sample size of 1700 was reached. 732/1700 (43.06%) of the participants were male, while the remaining 968 (56.94%) were female. **Table 3** below shows the number of people recruited from each district. 264 (15.53%) were children <10 years, 489 (28.75%) were between the ages of 10 and 19 years and 947 (55.71%) were 20 years and above. The ages of the participants ranged from 5 to 100 years, with a median of 23 years and a mean age of 28 years. From the participants, skin snips (2) [from adults only], DBS (3), plasma (3), whole blood (2) aliquots were collected and stored to establish a biobank of samples for future evaluations and assessments.

### RDT results on whole blood in the field

Of the 1700 participants, four (4) refused blood draw. Thus, 1696 participants were tested with the BA/GADx and DDTDc RDTs in the field. 295 tests were positive with BA/GADx, while 374 tests (187 for T1&T2, 155 for T1 only and 32 for T2 only) were positive by the DDTDc RDT. **Tables 4 and 5** below show the breakdown in test positivity for the BA/GADx and DDTDc RDTs in the field, respectively. Invalid test results were only observed in the Adaklu district. The LF FTS was only performed in the Adaklu district, with 2/656 (0.68%) people testing positive. No *Wuchereria bancrofti* microfilariae were detected in the night blood collected.

### Infection prevalence based on the SD Ov16, DDTDc and BA/GADx RDTs

Of the 1696 participants tested with the BA/GADx and DDTDc RDTs in the field, 1672 agreed to blood collection and storage for later testing in the lab. Infection prevalence using the SD Ov16 RDT was 3.36%, 18.82% and 29.22% in the Adaklu, Hohoe and Nkwanta North districts, respectively (**Table 6**). When compared to the infection prevalence based on the new RDTs in the field and laboratories, the results were generally comparable between tests, with overlapping 95% confidence intervals (***Figure 1***). For all the tests, fewer positives were detected from the Adaklu district (considered non-endemic), with higher infection prevalence in the Hohoe and Nkwanta North districts. When analysed by age, the percentage positivity by the different tests increased gradually, reaching a peak in the 40-to-60-year age groups (***Figure 2***). There was a significant difference in the positive results between age groups with p ≤ 0.0003 for each test. There was substantial to almost perfect agreement between the different tests (See Supplementary File 2).

**Figure 1:**
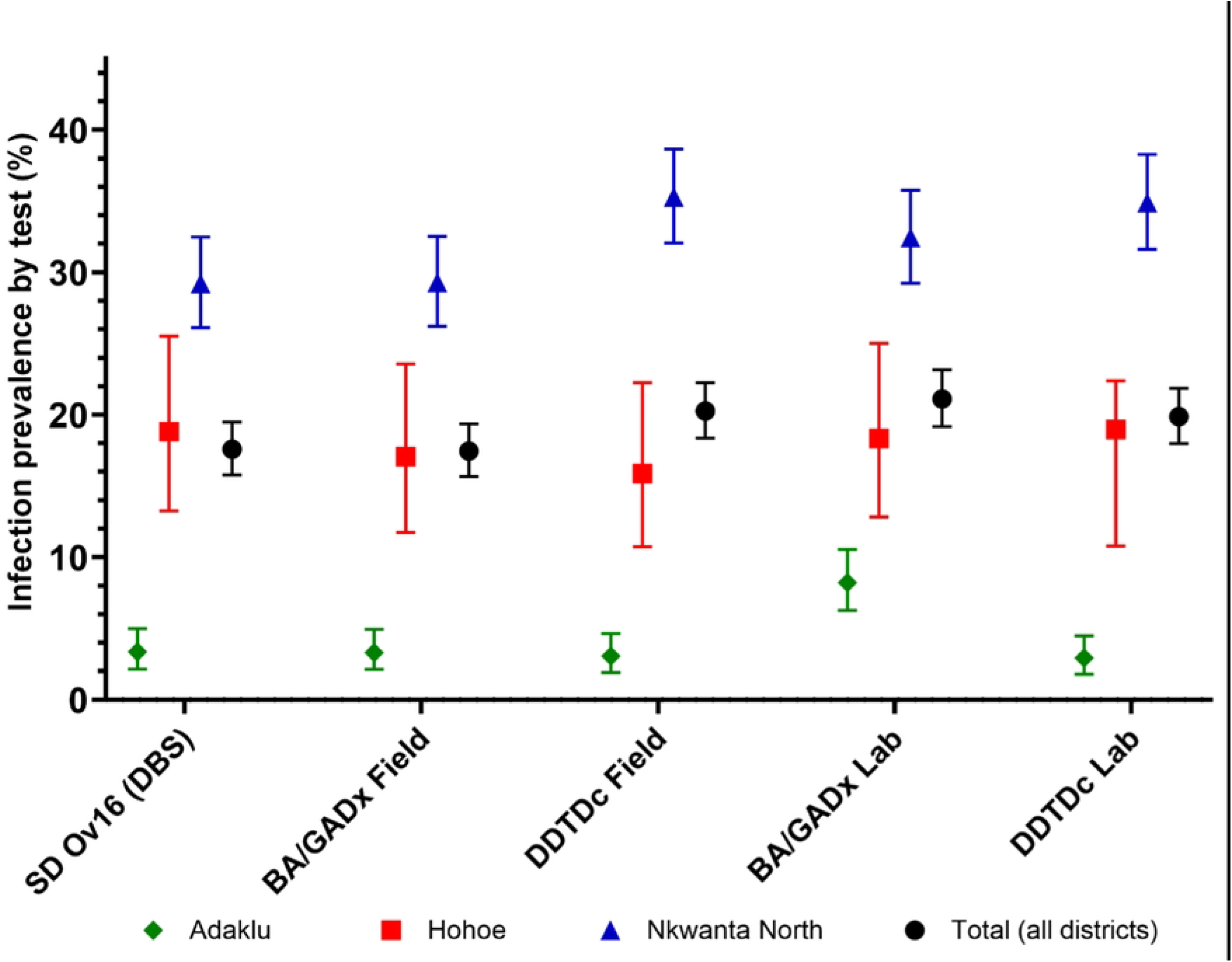
O. volvulus infection prevalence based on the SD Ov16 RDT in the lab, BA/GADx (field and lab) and DDTDc (field and lab) tests. Error bars represent the 95% Confidence Intervals.

**Figure 2:**
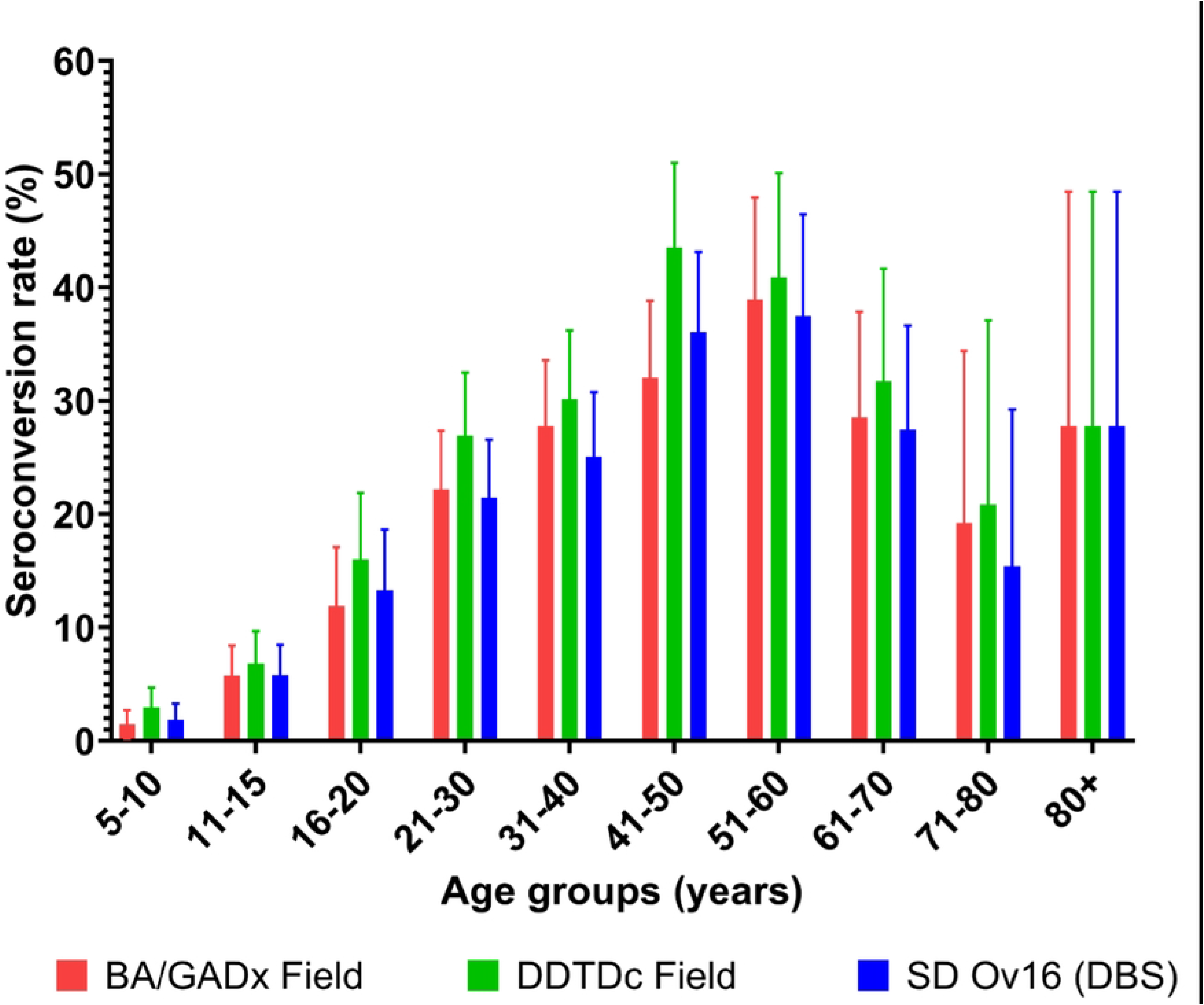
Seroconversion rates by age group. Error bars represent the 95% Confidence Intervals.

### Sensitivity of the field and laboratory DDTDc tests compared to the SD Ov16 RDT

The concordance between the field DDTDc (tests positive for T1, irrespective of T2 results) and SD Ov16 RDT laboratory tests was 92.40%. The Kappa value was 0.632 (SE: 0.032: 95% CI: 0.570 to 0.694) indicating substantial agreement between the two tests. The sensitivity of the DDTDc test on whole blood in the field, compared to the SD Ov16 on eluted DBS was 78.52% (95% CI: 71.06 to 84.82) (***Figure 3***). The concordance between the laboratory DDTDc (tests positive for T1) on eluted DBS and SD Ov16 RDT on eluted DBS was 92.56%. The Kappa value was 0.755 (SE: 0.021: 95% CI: 0.714 to 0.796) indicating substantial agreement between the two tests. The sensitivity of the DDTDc test on eluted DBS, compared to the SD Ov16 on eluted DBS was 85.57% (95% CI: 81.00 to 89.40). When considering the test as positive only when both T1 and T2 are present, the sensitivity of the DDTDc test on whole blood in the field, compared to the SD Ov16 on eluted DBS was 49.32% (95% CI: 43.47 – 55.19). The sensitivity of the test on eluted DBS in the laboratory, compared to the SD Ov16 was 45.52% (95% CI: 39.69 – 51.44).

**Figure 3:**
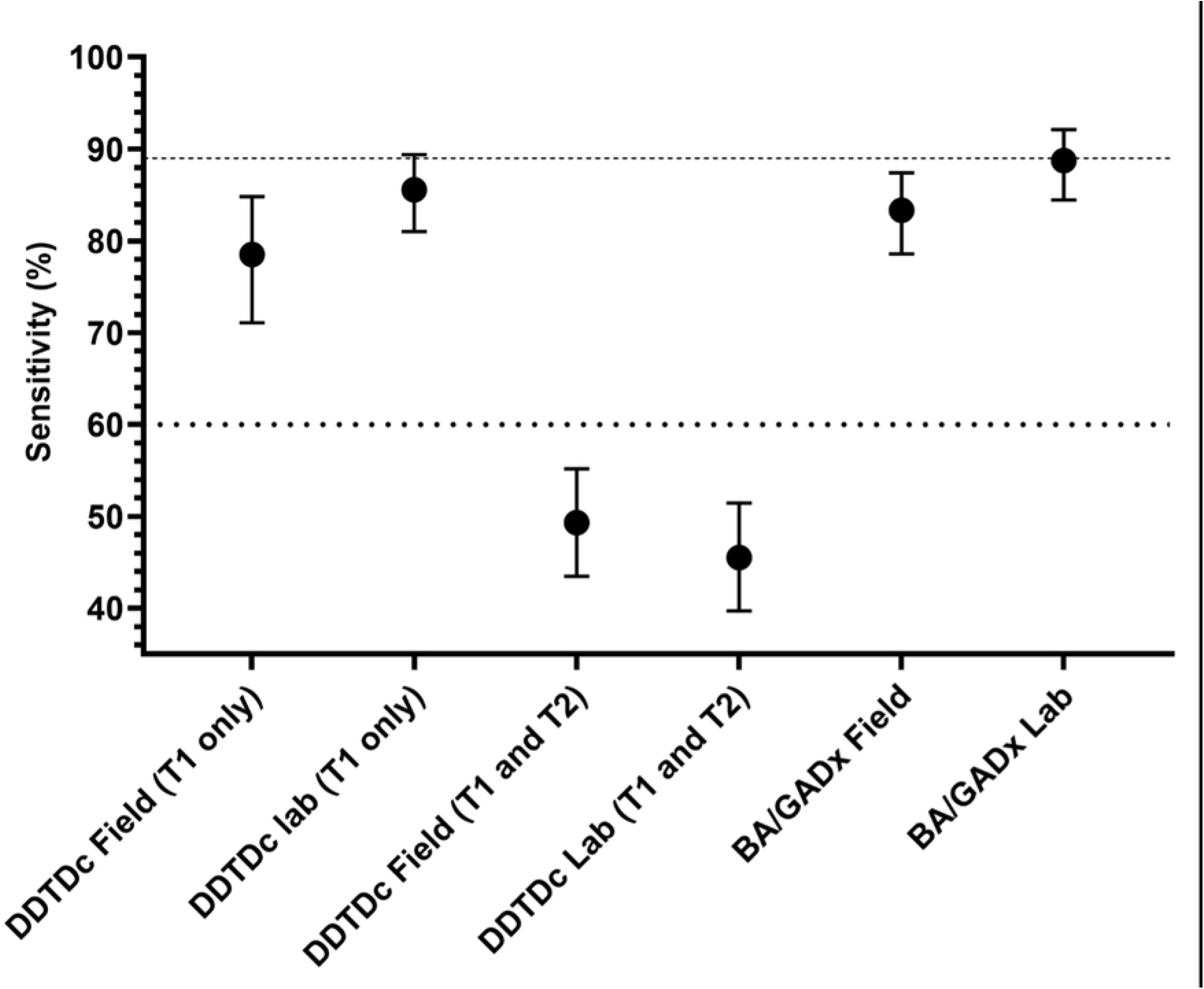
Sensitivity of tests using the SD Ov16 RDT as reference. Error bars represent the 95% Confidence Intervals. The dotted line at the bottom is the WHO TPP sensitivity for RDTs to be used for mapping decisions, while the dashed line at the top is the WHO TPP sensitivity for stopping decisions.

### Sensitivity of the field and laboratory BA/GADx tests compared to the SD Ov16 RDT

The concordance between the field BA/GADx test on whole blood and SD Ov16 RDT test on eluted DBS was 94.01%. The Kappa value was 0.794 (SE: 0.020: 95% CI: 0.755 to 0.833) indicating a substantial agreement between the two tests. The sensitivity of the BA/GADx test on whole blood in the field, compared to the SD Ov16 on eluted DBS was 83.33% (95% CI: 78.57 to 87.41) (***Figure 3***). The concordance between the laboratory BA/GADx test on eluted DBS and SD Ov16 RDT test on eluted DBS was 92.50%. The Kappa value was 0.760 (SE: 0.020: 95% CI: 0.720 to 0.800) indicating a substantial agreement between the two tests. The sensitivity of the BA/GADX test on DBS, compared to the SD Ov16 on eluted DBS was 88.74% (95% CI: 84.55 to 92.12).

### Comparison between the sensitivity of the two tests using the SD Ov16 as the reference

Overall, sensitivity of the BA/GADx test was slightly higher than the DDTDc test (***Figure 3***), but confidence intervals overlap. For both RDTs, the tests run on eluted DBS in the laboratory provided the best results in terms of sensitivity.

### Skin snip microscopy and O-150 qPCR

Skin snip microscopy was conducted for 907 adults. 18 samples were positive (1.98%; 95% CI: 1.18 – 3.12). By district, the positivity was 0% in Adaklu, 4.90% (5/102 positives; 95% CI: 1.61–11.07) in Hohoe, and 2.83% (13/459 positives; 95% CI: 1.52–4.79) in Nkwanta North.

Of these samples, 26 skin snip samples tested positive using the O-150 qPCR [18], all from Nkwanta North (5.66%; 95% CI: 3.73 – 8.19). Only five (5) samples were positive by both microscopy and qPCR, whereas 21 samples were positive by qPCR and negative for microscopy, and 13 samples were positive by microscopy and negative for qPCR.

### Sensitivity of the field and laboratory DDTDc tests compared to the skin snip qPCR

Looking at any tests positive for antibodies against Ov16, the concordance between the field DDTDc test on whole blood and skin snip qPCR was 68.52%. The Kappa value was 0.075 (SE: 0.019: 95% CI: 0.037 to 0.113) indicating a slight agreement between the two tests. The sensitivity of the DDTDc test on whole blood in the field, compared to the skin snip qPCR was 76.9% (95% CI: 56.4 to 91.0). When compared to the laboratory results, the concordance between the laboratory DDTDc test on eluted DBS and skin snip qPCR was 69.53%. The Kappa value was 0.067 (SE: 0.020: 95% CI: 0.028 to 0.106) indicating a slight agreement between the two tests. The sensitivity of the laboratory DDTDc test, compared to the skin snip qPCR was 69.2% (95% CI: 48.2 to 85.7). When considering the test as positive only when both T1 and T2 are present, the sensitivity of the DDTDc test on whole blood in the field, compared to the skin snip qPCR was 61.54% (95% CI: 40.57 - 79.77). The sensitivity of the laboratory DDTDc test, compared to the skin snip qPCR was 53.85% (95% CI: 33.37 - 73.41).

### Sensitivity of the field and laboratory BA/GADx tests compared to the skin snip qPCR

The concordance between the field BA/GADx test on whole blood and skin snip qPCR was 72.48%. The Kappa value was 0.085 (SE: 0.022: 95% CI: 0.041 to 0.129) indicating a slight agreement between the two tests. The sensitivity of the BA/GADx test on whole blood in the field, compared to the skin snip qPCR was 73.1% (95% CI: 52.2 to 88.4). Compared to the laboratory results, the concordance between the laboratory BA/GADx test on eluted DBS and skin snip qPCR was 68.22%. The Kappa value was 0.062 (SE: 0.019: 95% CI: 0.025 to 0.099) indicating a slight agreement between the two tests. The sensitivity of the laboratory BA/GADx test, compared to the skin snip qPCR was 69.2% (95% CI: 48.2 to 85.7).

### Sensitivity of the laboratory SD Ov16 test compared to the skin snip qPCR

The concordance between the laboratory SD Ov16 test on eluted DBS and skin snip qPCR was 72.70%. The Kappa value was 0.073 (SE: 0.022: 95% CI: 0.029 to 0.116) indicating a slight agreement between the two tests. The sensitivity of the laboratory SD Ov16 test, compared to the skin snip qPCR was 65.4% (95% CI: 44.3 to 82.8).

### Specificity and non-inferiority assessments

Due to the limitations in the low number of skin snip positives detected, specificity and non-inferiority assessments of the tests could not be undertaken using the O-150 qPCR as a reference.

### Mansonella qPCR

Of the 1673 samples analyzed, 78 samples were positive by the Mansonella qPCR. Table 7 shows the result of the *Mansonella perstans* qPCR. The positivity between sites ranged from 0.61% to 25.73%, with high positivity detected in the Hohoe district. Of the 78 *M. perstans* positives, 22 were positive for DDTDc T1 in the laboratory, 29 were positive for BA/GADx in the laboratory, 20 samples were positive for DDTDc T1 in the field, and 25 were positive for BA/GADx in the field.

### Feasibility and Ease of Use

Having performed more than 3400 of each test, four test readers completed an ease-of-use questionnaire. All found the tests to be feasible, acceptable, and easy to use in the field and laboratory. However, for the BA/GADx test three of the readers indicated that the presence of a blood smudge on the test after running, which made it difficult to correctly read the results. For the DDTDc test, one reader reported the difficulty in reading very faint test lines.

## Discussion

The new RDTs were field-friendly, easy to use, and comparable to the SD Bioline RDT, making them practical for onchocerciasis programmes, except for the need for some optimizations. Importantly, they can be used in the field at the point of care, which is an advantage over the SD Ov16 RDT. The SD Bioline RDT lacks sensitivity when used with blood collected directly from a fingerstick and is only sensitive enough for program use when run in the laboratory with DBS.

When looking at the Ov16 signal alone and using the SD Ov16 RDT as the reference, the BA/GADx test demonstrated slightly higher sensitivity (Field: 83.33%; DBS: 88.74%) compared to the DDTDc test (Field: 78.52%; DBS: 85.57%), with overlapping confidence intervals. Overall, there was a slight improvement in the sensitivity of the tests in the laboratory compared to the field, suggesting that factors such as environmental conditions, operator handling, and sample matrices (whole blood vs DBS) may account for the difference in the performance of these RDTs.

Given the two test lines on the DDTDc test we can analyse results further, particularly in the comparison of positive definition: the presence of T1 as positive aligns more closely to the currently accepted RDT or while the manufacturer recommends interpreting tests as positive only when both T1 and T2 are present. As T1 detects antibodies to Ov16, similar to the BA/GADx and SD Bioline tests, it is unsurprising that similar sensitivity and specificity were observed when comparing these two tests. However, considering the DDTDc as positive only when both test lines are present leads to a significant decline in sensitivity and improvements in specificity. This can be explained by the fact that, unlike Ov16, OvOC3261 is expressed only after appearances of the microfilariae [21]. Thus, it may be valuable to utilize the DDTDc test in different use cases depending on the testing scenarios. In mapping scenarios, where the prevalence of the disease is high, considering any test positive when T1 is present may be acceptable when high sensitivity is required. However, in stop-MDA settings, where it is important to ascertain ongoing transmission, considering the test as positive only when both T1 and T2 are present leads to the required high-test specificity. However, it will be important for these use cases to be more thoroughly defined within the onchocerciasis community in order to allow the use of these tests. Usability of a test with multiple test lines must be taken into consideration when considering implementation options.

The study demonstrated that concordance between the BA/GADx and DDTDc tests was relatively high, with agreement levels above 89% in both field and laboratory settings. The kappa values further indicated substantial agreement between the two tests. However, when compared to traditional diagnostic methods such as skin snip microscopy and O-150 qPCR, the agreement was lower. This is expected, as antibody-based tests such as the BA/GADx and DDTDc RDTs primarily detect past or current exposure rather than active infection, whereas qPCR and skin snip microscopy are more indicative of ongoing parasitemia, which was detected at very low prevalence in the study communities.

The discordance between the microscopy and the qPCR results is worth noting. This may point to the sensitivity and specificity differences between the two tests. On one hand, microscopy coupled with the ability of the microscopists, may not be sensitive enough to identify very low levels of infections [22]. By contrast, the O150 qPCR assay may be prone to suboptimal primer/probe binding resulting in decreased specificity [18, 23]. While residual skin snip biopsies and emerged microfilariae were transferred in eppendorf tubes after microscopy, it is possible that some microfilariae may have been missed during the transfer, and hence qPCR results could be negative despite a positive *O. volvulus* result by microscopy. Further, the co-endemicity of *O. volvulus* and other filarial parasites may lead to false positives by microscopy or PCR methods. *Mansonella streptocerca* microfilariae do manifest in the skin and may thus confound the microscopy results [24, 25]. Preliminary data from sequenced DNA extracted from skin snips in this study reveal the presence of *M. streptocerca* in some samples (Unpublished). *M. Streptocerca* has been previously reported in Ghana [26].

The discordance observed between RDT results and skin snip qPCR suggests that molecular methods remain the most accurate approach for confirming active infections. This aligns with previous studies that have emphasized the limitations of antibody-based tests for determining infection status in endemic populations [22, 27]. Given that qPCR detected cases that were missed by microscopy, its utility as a confirmatory tool should be further explored in elimination settings.

The introduction of new, more sensitive diagnostic tools without need of laboratory infrastructure is essential for accelerating onchocerciasis elimination efforts, particularly their use in hypo-endemic areas and in post-treatment surveillance [3]. The results from this study suggest that while the BA/GADx and DDTDc tests provide useful diagnostic options, their interpretation in programmatic settings requires careful consideration. The ability of these tests to detect exposure to *O. volvulus* could aid in mapping areas of transmission and identifying areas where continued MDA is necessary. However, because these RDTs detect antibodies rather than active infection, they may be less suitable for making decisions regarding stopping MDA.

A couple of limitations were identified for this study. First, the study was conducted in a limited number of sites within Ghana, and results may not be generalizable to all onchocerciasis-endemic regions. Second, the skin snip microscopy and qPCR resulted in very low numbers detected, leading to the inability to effectively use these in the assessment of the diagnostic performance of the new RDTs. While the sensitivity of the new tests was assessed using the SD Ov16 RDT, any comparisons with the WHO TPP characteristics (clinical sensitivity requirements for mapping (>60%) and for stopping treatment (>89%)) should be interpreted with care, as the TPP characteristics are to be compared with the qPCR results, which unfortunately, were too limited for any meaningful analyses, and non-inferiority comparison in this study. Finally, new versions of these tests have been developed, the performance of which may need reassessment to confirm their performance.

Overall, the findings from this study indicate that both the BA/GADx and DDTDc RDTs offer promising alternatives to the existing diagnostic tools for onchocerciasis. Their rapid, point-of-care applicability makes them valuable for field settings where access to laboratory facilities is limited. However, their use should be complemented by confirmatory molecular diagnostics to improve accuracy, particularly in settings approaching elimination. Further evaluations in different epidemiological contexts will be necessary to determine their long-term utility in onchocerciasis control and elimination programs.

## Acknowledgements

We are grateful to the community members and the staff of the different District Health Management Teams for their support during this study. We are grateful to Mr. Daniel Adjei Odumang, Ms. Bertha Klenam Hamenu, Mr. Joseph K. Quartey, Mr. Kojo Nketiah, Ms. Enuenyam Naana Ninson; Ms. Janet Abena Ampofo, Ms. Ruth Adjaloko, Ms. Grace Gyemfi, Mr. Sampson Otoo, Mr. Dickson Osabutey, Mr. Isaac Quaye, Ms. Thelma Koo, Mr. Christopher Dorcoo and Mr. Edward Dumashie, all of whom supported with the field data collection.

## Author contributions

Design: **DKdS, LH, KYW, CV, PL, MB, DAB**; Methodology: **LH, PK, JP-G, SEE, KYW, CV, PL, AD, EA, MB**.

Data collection and validation: **DKdS, YA, SA, MD, AAN, BL, KF**; Data analysis: **DKdS, CM**; Data curation: **BL**; Writing—original draft preparation: **DKdS, YA, SA, MD, AAN**; Writing—review and editing: **KF, CM, PK, JP-G, SEE, KYW, CV, PL, AD, EA, MB, DAB**; Visualization: **DKdS;** Project administration: **DKdS**; Funding acquisition: **DKdS**. All authors have read and agreed to the published version of the manuscript.

## Data availability statement (mandatory)

All data generated or analysed during this study are included in this published article (and its Supplementary Information files). The raw data has been submitted to the WHO DTAG subgroup for onchocerciasis for collation and analysis together with the data from other countries. The raw data will be made available on the website of the Coalition for Operational Research on Neglected Tropical Diseases after publication. A request for the data can be made to Carson Moore.

## Disclaimer

The findings and conclusions in this paper are those of the authors and do not necessarily represent the official position of the Centers for Disease Control and Prevention, nor do they suggest endorsement of specific products or trade names used in this study.

## Tables

- Table 1: Study sites identified for the evaluation.
- Table 2: Summary table for sample collection and processing. Note: some participants refused venous blood collection. Thus, all laboratory processes for such participants were not conducted
- Table 3: Number of participants recruited from each district.
- Table 4: Test results based on the BA/GADx RDT on whole blood in the field.
- Table 5: Test results based on the DDTDc RDT on whole blood in the field.
- Table 6: *O. volvulus* infection prevalence based on the SD Ov16 RDT.
- Table 7: Prevalence of *M. perstans* in the study areas.

## Supplementary files (S)

S1. Ease-of-use questionnaire

S2. Concordance between test results

